# T-cell and antibody immunity after COVID-19 mRNA vaccines in healthy and immunocompromised subjects-An exploratory study

**DOI:** 10.1101/2021.05.21.21257442

**Authors:** Rakesh Sindhi, Chethan Ashokkumar, Brianna Spishock, Maggie Saunders, Angelo Mabasa, Pradeep Sethi, Ashok Reddy, Bobby Nibhanupudy

**Affiliations:** University of Pittsburgh; Plexision Inc, Pittsburgh; Columbia University Medical Center, NY; AdventHealth, Orlando, FL

## Abstract

**Background:** In recent studies, up to half of immunocompromised (IC) subject populations fail to develop antibodies after COVID-19 vaccination.

**Purpose and Methods:** Here, we explore whether T-cells which respond to the spike (S) antigenic sequence and its less conserved S1, and the conserved S2 component are present in serial samples before and after each dose of mRNA1273 or BNT162b2 vaccines in 20 healthy immunocompetent subjects. Single samples from 7 vaccinated IC subjects were also tested. Simultaneously, we measured IgG antibodies to the receptor binding domain (RBD) of S1, and anti-S IgG, and frequencies of monocytic CD14+HLA-DR-(M-MDSC) and polymorphonuclear CD14-CD15+CD11b+ (PMN-MDSC) myeloid-derived suppressor cells.

**Results:** In healthy subjects, S1-, S2-, and S-reactive CD4 and CD8 T-cell frequencies showed a numeric but not statistically significant decrease after the first vaccine dose and were accompanied by increased MDSC frequencies (p<0.05). After the second dose, S2-and S-reactive CD4 and CD8 cells and MDSC approached pre-vaccination levels. In healthy subjects, a) S1-reactive CD8 frequencies were significantly higher after the second dose compared with pre-vaccination levels (p=0.015), b) anti-RBD and anti-S IgG were present in all after the second dose. Among seven IC subjects, anti-RBD and anti-S IgG were absent in 4 and 3 subjects, respectively. S1-reactive CD8 cells were identified in 2 of 4 anti-RBD negative subjects. S-reactive CD4 or CD8 cells were identified in all three anti-S negative subjects.

**Conclusions:** In healthy immunocompetent subjects, mRNA vaccines induce antibodies to the spike antigenic sequences and augment CD8 cells reactive to the S1 spike sequence, which is more specific for the SARS-CoV-2 virus. In this exploratory cohort of vaccinated immunocompromised subjects, S1-reactive CD8 cells can be detected in some who are negative for RBD antibody, and S-reactive T-cells are present in all who are negative for spike antibody.

## Introduction

Cellular immunity assessments are needed after COVID-19 vaccination because antibodies do not develop in all vaccinated immunocompromised (IC) individuals. To illustrate, only 15% of transplant patients receiving chronic immunosuppression were found to have an antibody response after the first dose of mRNA vaccines^1^. This number increased to 54% after the second dose, far lower than what has been reported from healthy immune competent subjects^2,3^. In a study of inflammatory bowel disease patients receiving the anti-tumor necrosis factor antibody infliximab, vaccination was more likely to produce an antibody response after the second dose than the first dose of COVID-19 vaccines^4^. Nearly half of all patients with hematologic malignancies also failed to make antibodies after COVID-19 vaccination^5^. Among those with B-cell malignancies, the number decreased to 23%. It is not known whether T-cell or other cellular immune responses can compensate for failure to develop antibodies in IC patients. To address this question, cellular and humoral immunity must also be assessed contemporaneously in healthy subjects using robust assays.

We have recently described decreased spike-antigen-reactive T-cells and IgG antibodies to the receptor binding domain (RBD) of the spike protein early after COVID-19 infection in chronically immunosuppressed transplant patients^6^. The S antigenic sequence is comprised of the less conserved N-terminal S1 component, which contains the receptor binding domain (RBD), and the conserved C-terminal S2 sequence^7^. The T-cell assay uses CD154, which is co-expressed with interferon gamma (IFNγ) upon S antigen stimulation, as the single marker of viral-antigen-specific T-cells^6,8^. Assay reproducibility is reflected by a coefficient of variation ranging from 2-10.6% under a variety of conditions^6^. In our recent study, Impaired T-cell responses to S antigen were accompanied by an increase in circulating myeloid derived suppressor cells (MDSC) early after COVID-19 infection^6^. These cells arise during acute viral infections and suppress T-cells^9^.

In this exploratory study, we characterize T-cell and IgG responses to S antigenic sequences before and after each dose of the mRNA vaccines in 20 healthy immunocompetent subjects. Single samples from seven vaccinated IC subjects are also evaluated with the same assays.

## Results

Study protocols were reviewed and approved by Advarra Institutional Review Board (IRB), Columbia, MD. Subjects were enrolled under IRB-approved protocols Pro00045352 and Pro00053511 (Advarra IRB, Columbia, MD) (NCT#04883164). Informed consents were obtained from all participants in the study. For 20 healthy individuals, mean (SEM) age was 51 ± 3.6 years, male: female distribution was 8: 12 and race Caucasian: African-American: Asian was 15: 1: 4. Eleven subjects received mRNA1273 (Moderna, Cambridge, MA) and 8 received BNT162b2 (Pfizer, New York, NY). For 7 IC subjects, mean age was 45 ± 9 years. Subjects included 4 males and 3 females, 5 Caucasians, 1 African-American and 1 Asian. Six subjects received BNT162b2 and one received mRNA1273. Four received both doses, and three received a single dose of the vaccine. The population included three solid organ transplant recipients (SOT), two with autoimmune disorder, one-bone marrow transplant recipient, and one with hematologic malignancy. IC recipients were on a variety of immunosuppressive medications, which included tacrolimus, belatacept, steroids, velcade, cellcept, leflunomide, and dimethyl fumarate,

### S1-reactive CD8 cells increase after vaccination in healthy immunocompetent subjects

PBL from each healthy subject, obtained pre-vaccination and at 2 weeks after each vaccination dose from healthy subjects were stimulated overnight with overlapping peptides mixtures representing the S1, S2 and S antigenic sequences as described previously^6^ (JPT Peptides, Germany). The frequencies of CD3, CD4, and CD8 cells that expressed CD154 after overnight stimulation with S antigenic peptides were measured with flow cytometry as described previously^6^. These frequencies were also measured in single samples obtained from 7 IC subjects. Monocytic (M-MDSC) and polymorphonuclear (PMN-MDSC) MDSC frequencies were measured as described previously^6^.

S2-and S-reactive CD4 and CD8 cells showed a numerical but not statistically significant decrease after the first vaccine dose and were reconstituted to pre-vaccination levels after the second (Figure 1, Table 1). An exception is frequencies of S1-reactive CD8 cells which were significantly higher after the second dose compared with pre-vaccination frequencies (p=0.015). This pattern is mirrored in reverse by M-MDSC and PMN-MDSC, which increased significantly after the first dose. Mean±SEM frequencies (%) of M-MDSC were 4.7±1.1 pre-vaccination, increased significantly to 28.3±5.5, p=0.004, after the first dose, and decreased but remained significantly elevated at 13.9±1.9, p=0.002, after the second dose compared with pre-vaccination levels (Figure 1). PMN-MDSC frequencies were 9.2±1.6 pre-vaccination, increased significantly to 18.6±3.3, p=0.019, after the first dose, and returned to pre-vaccination levels 11.5±1.5, p=0.31, NS..

**Table 1.** Mean ± SEM frequencies (%) of S1-, S2- and S-reactive CD4 and CD8 cells, and OD_490_ for anti-RBD and anti-S IgG in 20 healthy immunocompetent subjects in samples obtained pre-vaccination, and 2 weeks after the first and the second doses of mRNA vaccines. Also shown are corresponding measurements in single samples from 7 immunocompromised (IC) subjects. * indicates significant difference compared with pre-vaccination levels (p=0.015)

|  | CD4 |  |  | CD8 |  |  | RBD IgG | SPIKE IgG |
| --- | --- | --- | --- | --- | --- | --- | --- | --- |
|  | S1 (%) | S2 (%) | S (%) | S1 (%) | S2 (%) | S (%) | OD490 | OD490 |
| Healthy-Pre vaccination (n=20) | 0.1±0.1 | 2.1±0.6 | 2.7±0.6 | 0.2±0.1 | 2.7±0.6 | 4.0±0.7 | 0.2±0.1 | 0.2±0.1 |
| Healthy-First dose (n=20) | 0.1±0.0 | 1.5±0.5 | 1.6±0.3 | 0.2±0.1 | 2.2±0.7 | 2.1±0.4 | 1.6±0.3 | 1.9±0.2 |
| Healthy Second dose (n=20) | 0.1±0.1 | 1.9±0.3 | 2.1±0.3 | 0.7±0.2* | 3.1±0.6 | 3.1±0.5 | 2.8±0.1 | 2.9±0.1 |
| IC (n=7) | 0.6±0.3 | 1.8±0.5 | 1.1±0.3 | 0.7±0.5 | 1.9±0.7 | 2.0±0.7 | 1.0±0.5 | 1.3±0.5 |

**Figure 1.**
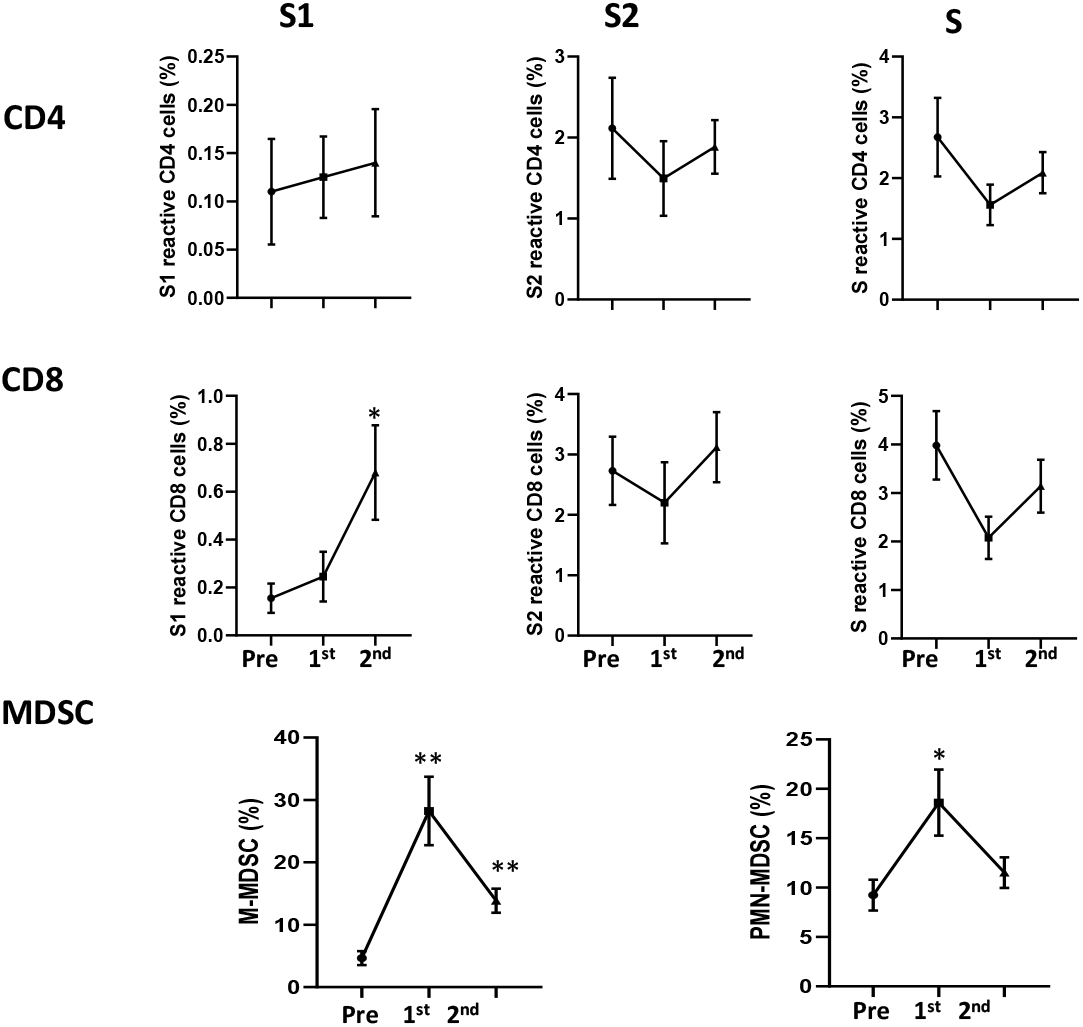
Line diagrams show mean ± SEM frequencies of S1- S2- and S-reactive CD4 cells (upper panel) and CD8 cells (middle panel) pre-vaccination (pre) after the first dose (1^st^) and second dose (2^nd^) of mRNA vaccines. At each timepoint, SEM are plotted as error bars. Lower panel shows changes in frequencies of monocytic (M-MDSC) and polymorphonuclear MDSC (PMN-MDSC) at corresponding timepoints. * significant differences compared with pre-vaccination frequencies. * p<0.05, ** p<0.005.

### Antibody incidence and strength increases after vaccination in healthy immunocompetent subjects

Anti-RBD and anti-S IgG were measured with ELISA as the optical density at 490 nm (OD_490_). An OD490 of 0.45 or greater implied a positive test for each antibody as described previously^6^. The incidence of anti-RBD and anti-spike IgG increased from 17/20 and 18/20 two weeks after the first dose, to 20/20 and 20/20 two weeks after the second dose (Figure 2). Consistent with this trend, the mean ± SEM OD_490_ for anti-RBD IgG and anti-S IgG increased progressively after each dose compared with pre-vaccination measurements. These OD_490_ values were noteworthy for the larger interindividual variation represented by higher SEM values after the first vaccine dose, and smaller variation after the second dose.

**Figure 2.**
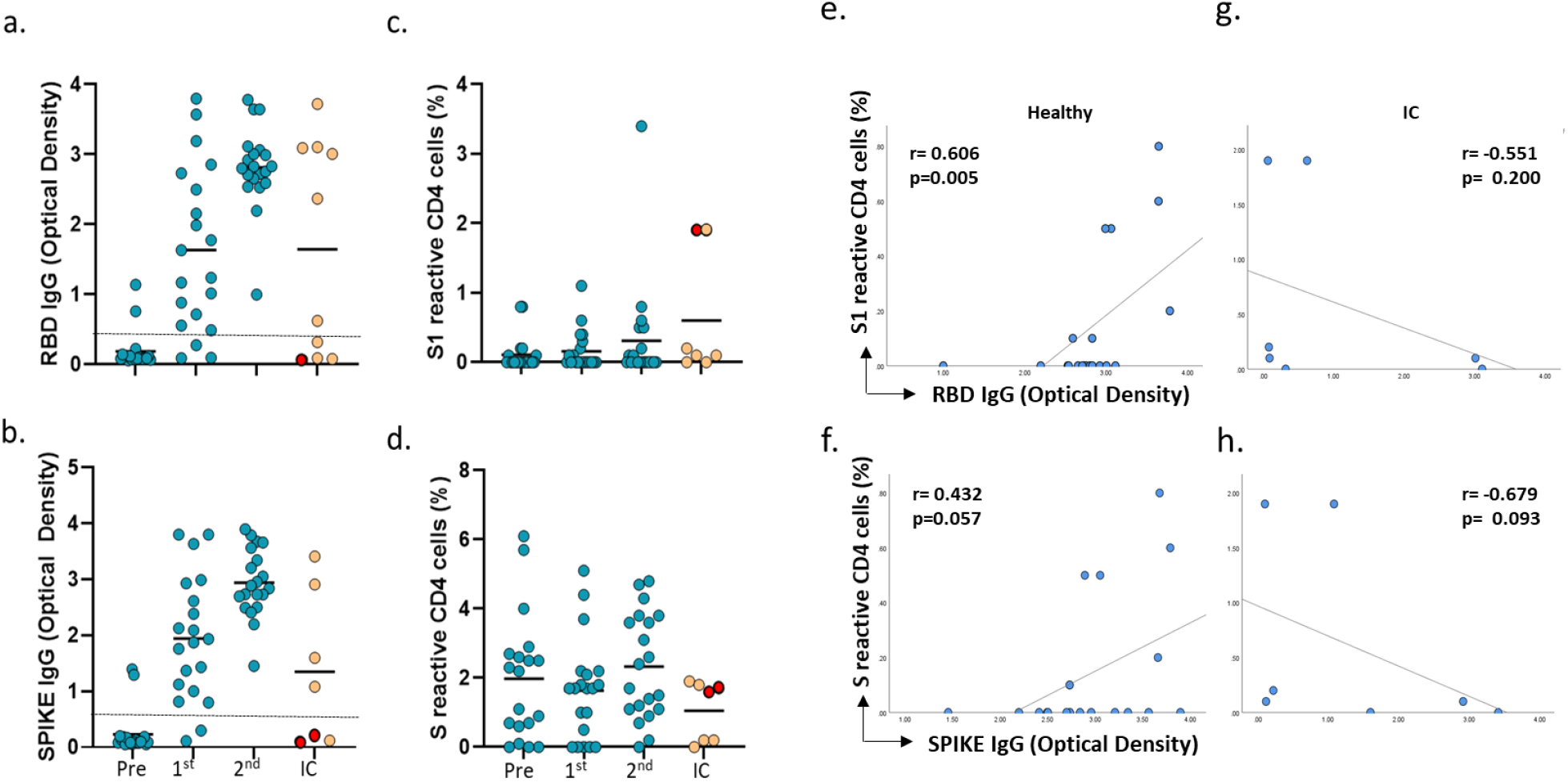
Optical density at 490 nm (OD_490_) for **(a)** Anti-RBD IgG and **(b)** Anti-spike IgG in 20 healthy immunocompetent subjects pre vaccination (Pre), after the first (1^st^) and the second vaccine doses (2^nd^) in 20 healthy immunocompetent subjects. Orange circles show OD_490_ for each antibody in single samples after the first or second dose from 7 immunocompromised (IC) subjects. Dotted lines in (a) and (b) indicates OD_490_ of 0.45 at or above which the antibody test is positive. **(c)** S1-reactive CD154+CD4 T-cell frequencies in healthy and IC subjects. **(d)** S-reactive CD154+CD4 T-cell frequencies in healthy and IC subjects. IC subjects who failed to make antibodies but had higher S1-reactive or S-reactive CD4 frequencies are identified with red circles. Spearman correlation plots show relationships between S1-reactive CD4 cells and anti-RBD IgG (e and g) and anti-spike IgG (f and h) in healthy (e and f) and immunocompromised subjects (g and h).

### Failure of antibody response is associated with S1-reactive or S-reactive T-cells in some IC subjects

Among IC subjects, anti-RBD and anti-spike IgG were present in 3/7 and 4/7 subjects based on OD_490_ of 0.45 or greater in these subjects. The mean ± SEM OD_490_ for anti-RBD and anti-S IgG were 1.05 ± 0.5 and 1.3 ± 0.5, respectively. Four IC subjects were negative for anti-RBD IgG, and three IC subjects were negative for anti-S IgG (Figure 2, Table 2). Two of four IC subjects who were negative for anti-RBD IgG were found to have S1-reactive CD8 cells that were in the upper half of the range of frequencies measured among IC subjects (Figure 3, Table 2). One of these subjects also demonstrated S1-reactive CD4 cell frequencies in the upper range (Figure 2, Table 2). These higher frequencies also approximated or exceeded mean frequencies of corresponding cells observed in 20 healthy immune competent subjects who had received both vaccine doses.

**Table 2.** S1-reactive CD4 cell frequencies-CD4-S1 (%) and CD8 cell frequencies-CD8-S1 (%), S-reactive CD4 cell frequencies-CD4-S (%) and CD8 cell frequencies-CD8-S (%), status of IgG antibody to RBD (RBD IgG) and spike protein (Spike IgG) in immunocompromised (IC) subjects. Antibody status - implies absence or a negative test, + implies present or a positive test. Condition: HEMAT: Hematologic Malignancy, Autoimmune: Autoimmune Disorder, BMT: Bone Marrow Transplant, SOT: Solid Organ Transplant.

| ID | Condition | CD4-S1 (%) | CD8-S1 (%) | CD4-S (%) | CD8-S (%) | RBD IgG | Spike IgG |
| --- | --- | --- | --- | --- | --- | --- | --- |
| IC_1 | HEMAT | 1.9 | 3.6 | 0 | 2 | - | - |
| IC_2 | Autoimmune | 0 | 0 | 0.2 | 0.8 | - | + |
| IC_3 | Autoimmune | 0.1 | 0 | 1.9 | 5.5 | + | + |
| IC_4 | BMT | 0 | 0 | 0.2 | 0.2 | + | + |
| IC_5 | SOT | 0.2 | 1.3 | 1.7 | 3.6 | - | - |
| IC_6 | SOT | 0.1 | 0 | 1.6 | 0.3 | - | - |
| IC_7 | SOT | 1.9 | 0.2 | 1.8 | 1.6 | + | + |

**Figure 3.**
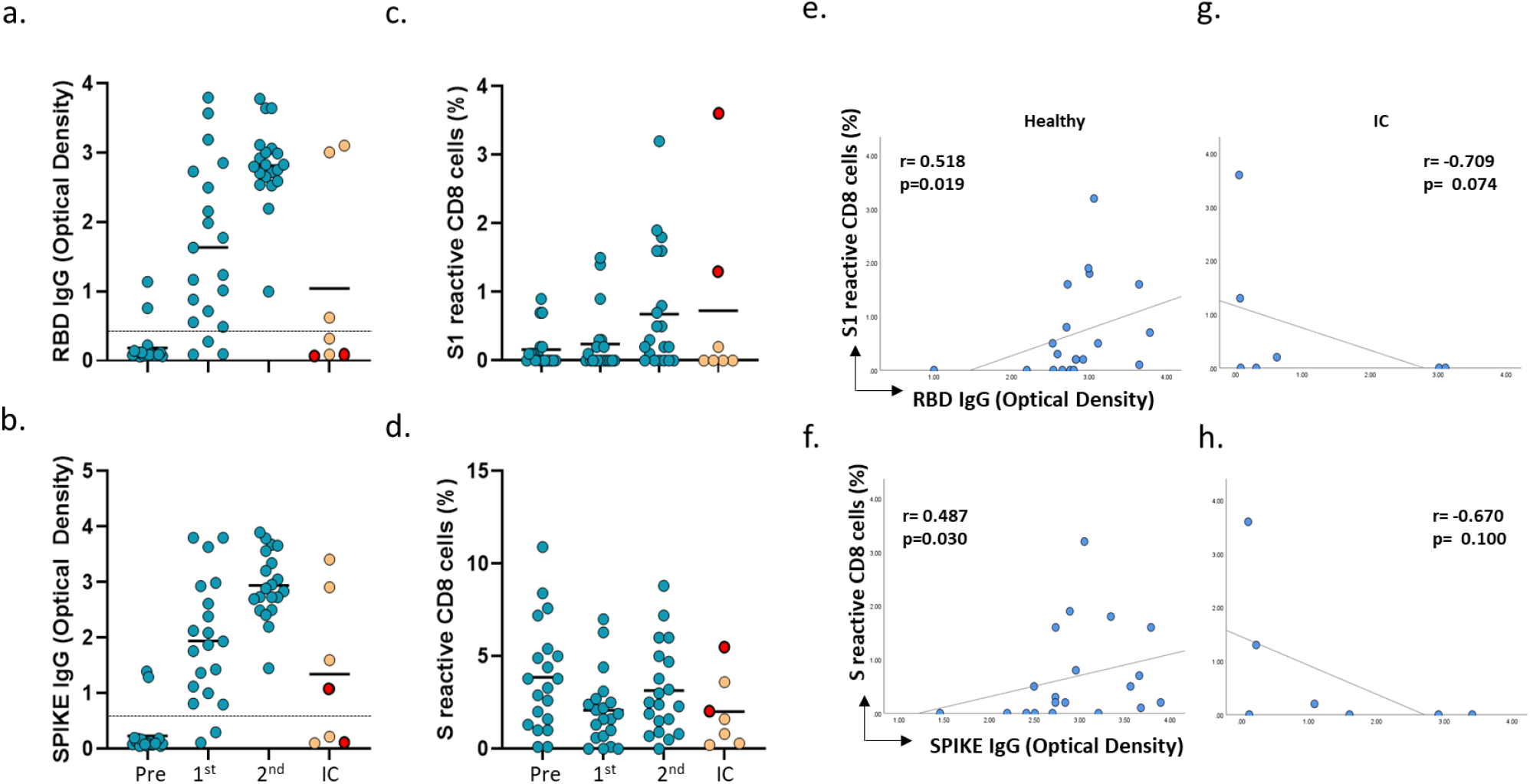
Optical density at 490 nm (OD_490_) for **(a)** Anti-RBD IgG and **(b)** Anti-spike IgG in 20 healthy immunocompetent subjects pre vaccination (Pre), after the first (1^st^) and the second vaccine doses (2^nd^) and in Immunocompromised (IC) subjects (orange circles) after the first or second vaccine dose. Dotted lines in (a) and (b) indicates OD_490_ of 0.45 at or above which the antibody test is positive. **(c)** S1-reactive CD154+CD8 T-cell frequencies in healthy and IC subjects. **(d)** S-reactive CD154+CD8T-cell frequencies in healthy and IC subjects. IC subjects who failed to make antibodies but had higher S1-reactive or S-reactive CD8 frequencies are identified with red circles. Spearman correlation plots show relationships between S1-reactive CD8 cells and anti-RBD (e and g) and anti-spike IgG (f and h) in healthy (e and f) and immunocompromised subjects (g and h).

All three IC subjects who were negative for anti-spike IgG were found to have S-reactive CD4 cells (Figure 2) or S-reactive CD8 cells (Figure 3) at frequencies that were in the upper half of the range for IC subjects. Among these three IC subjects, one subject each had either S-reactive CD4 or S-reactive CD8 cells at higher frequencies. Higher frequencies of both cell types were present in the third subject.

Low S1-reactive or S-reactive CD4 or CD8 frequencies of <1% were found in two IC subjects (Table 2). Both subjects expressed anti-spike IgG, with one also expressing anti-RBD IgG after vaccination

Multiple correlations were performed to evaluate relationships between OD_490_ of anti-RBD and anti-S IgG on the one hand and CD4 and CD8 cell frequencies that were reactive to the S1, S2 and S antigenic peptide mixtures on the other. For healthy vaccinated subjects, these correlations were sought between observations made after the second vaccine dose.

Among 20 healthy subjects, S1-reactive CD4 cells were positively correlated with the OD_490_ of anti-RBD IgG (r 0.61, p 0.005) and anti-S IgG (r 0.43, p 0.057). S1-reactive CD8 cells were also positively correlated with anti-RBD IgG (r 0.52, p 0.019) and anti-S IgG (r 0.49, p 0.03). Among the seven IC subjects, correlations between corresponding pairs of S1-reactive T-cells and the OD_490_ of anti-RBD and anti-spike IgG were negative but failed to achieve significance because of small numbers of patients in this group.

## Discussion

In healthy immunocompetent subjects, the first dose of mRNA vaccines appears to induce a state somewhat similar to that observed after less severe COVID-19 infection in our recent study^6^. CD4 and CD8 T-cells reactive to the various spike antigens showed a numeric decrease after the first vaccine dose (Figure 1). After the second dose, S2- and S-reactive CD4 and CD8 frequencies normalized to levels not statistically different from pre-vaccination frequencies. These changes in T-cells frequencies were accompanied by higher frequencies of M-MDSC and PMN-MDSC (p<0.05) after the first dose. After the second vaccine dose, M-MDSC frequencies approached but still exceeded pre-vaccination levels (p<0.05), while PMN-MDSC frequencies normalized to pre-vaccination levels. In our recent study, non-severe COVID-19 infection in which patients either had mild symptoms or were hospitalized without mechanical ventilation, S-reactive T-cell frequencies showed a numeric but not statistically significant decrease^6^. M-MDSC and PMN-MDSC increased early after COVID-19 infection. The inverse relationship between S-reactive T-cells and MDSC after COVID-19 infection was significant in these subjects with COVID-19 infection. MDSC are known to suppress T-cell function. High frequencies of MDSC after COVID-19 infection have also been reported by others^9^.

S1-reactive CD8 cells were the only T-cell subset for which frequencies increased significantly (p=0.015) after the second vaccination dose in healthy immunocompetent subjects, compared with pre-vaccination levels (Figure 1, Table 1). In contrast, the incidence and strength of anti-RBD and anti-spike IgG antibodies increased progressively after each dose of the vaccine (Figure 2). Antibody strength was inferred from OD_490_ values at each time point, pre-vaccination and after each dose. Interindividual variation for mean OD_490_ or SEM was larger after the first dose and smaller after the second dose. This finding suggested a more consistent level of antibody response after, and the importance of the second vaccine dose. We also observed positive correlations ranging from r values of 0.43 to 0.61 and p-values of 0.057 to 0.005 between S1-reactive CD4 cells and S1-reactive CD8 cells on the one hand, and the strength of anti-RBD and anti-spike IgG on the other hand, in healthy immunocompetent subjects (Figures 2 and 3).

Among vaccinated IC subjects, impaired antibody production and the presence of T-cells specific for SARS-CoV-2 may be related. Consistent with recent findings from vaccinated IC populations, anti-RBD and anti-spike IgG were present in 3 of 7 and 4 of 7 IC subjects and absent in 4 of 7 and 3 of 7 IC patients, respectively, in our study. However, two of these IC subjects who failed to make anti-RBD antibodies were found to have frequencies of S1-reactive CD8 cells (n=2) and S1-reactive CD4 cells (n=1) which approached or exceeded mean frequencies seen after complete vaccination in healthy immunocompetent subjects (Figures 3 and 2). All three IC subjects who failed to make anti-spike IgG showed higher frequencies of S-reactive CD4 or CD8 cells or both cell types. In all, 5 of 7 IC subjects were found to have S1- or S-reactive CD4 or CD8 cell frequencies >1%. The remaining two of seven IC subjects had low frequencies <1%, for each of the T-cells subsets that were reactive to the different spike antigens. Both these subjects were found to have anti-spike antibody, with one also expressing anti-RBD antibody.

Our inferences about the T-cell response to vaccination in IC subjects are limited by the fact that these subjects could not be sampled prior to vaccination. However, S1-reactive CD4 or CD8 cells reflect recent exposure to the virus or vaccine, because the S1 sequence has greater specificity for SARS-CoV-2 virus. Thus, the increase in S1-reactive CD8 cells after vaccination is suggestive of a post-vaccination augmentation in the T-cell response. This inference may be reasonable because S1-reactive CD8 cells were also the only T-cell subset to show a significant increase after the second dose in healthy immunocompetent subjects. This increase could compensate for failure to develop antibodies after vaccination in IC patients. Larger cohorts and functional studies are needed to confirm this possibility.

The clinical import of S2- and S-reactive T-cells is likely related to changes in MDSC and pre-existing immunity^10-13^. These cell types are indicative of prior exposure to structurally similar coronaviruses because the S2 sequence is relatively conserved among coronaviruses. S2- and S-reactive T-cells showed a transient and numeric but not statistically significant decline and were accompanied by a significant increase in MDSCs after the first vaccine dose in healthy immunocompetent subjects. MDSCs suppress T-cells. S2- and S-reactive T-cells normalized to pre-vaccination levels with MDSC also approaching pre-vaccination levels after the second dose. Because of the short follow-up after the second dose, the only reasonable inference is that pre-existing T-cell immunity is relatively unaffected after vaccination in healthy individuals. In IC subjects with a failed antibody response after vaccination, higher frequencies of S-reactive CD4 or CD8 cells may similarly imply pre-existing T-cell immunity which is unaffected by vaccination. Longer follow-up is needed to ascertain whether the early induction of MDSCs delays any potential augmentation of the T-cell response to conserved viral antigens.

The limitations of our exploratory study include the small numbers of immunocompromised vaccinated patients with different underlying conditions, our use of a single marker, CD154, in lieu of IFNγ as the marker of viral-antigen-specific cytotoxic T-cells, lack of longer-term serial immunity data from immunocompetent subjects, and single measurements from IC patients. However, our exploratory study shows that vaccination enhances T-cell immunity to SARS-CoV-2 in the form of S1-reactive CD154+CD8 T-cells in healthy immunocompetent subjects. Among IC subjects who failed to develop antibodies after vaccination, the presence of these S1-reactive CD8 cells is encouraging and warrants longitudinal confirmation in larger numbers of immunocompromised patients. Also worthy of additional study are S2- and S-reactive CD4 and CD8 cells, in vaccinated IC subjects, who do not develop anti-spike antibody. These studies can determine whether vaccination also augments pre-existing cellular immunity.

## Methods

Ficoll-purified peripheral blood leukocytes from each subject were stimulated with overlapping peptide mixtures (JPT Peptides, Germany) representing the S1, S2 and complete S antigenic sequences using non-permeabilizing staining as described^6^. Immunophenotyping of M-MDSC and PMN-MDSC was also performed as described^6^. All flurochrome-labeled antibodies were obtained from Biolegend (San Diego, CA) or BD Biosciences (San Jose, CA). IgG antibody to RBD and the spike protein S were measured with ELISA as described^6^. Statistical analysis and correlations were performed using SPSS. Dot plots were generated with GraphPad prism 9.

## Data Availability

All data that underlie the results reported in this Article (including study protocol) on individual participants will be made available to researchers who provide a methodologically sound proposal to the corresponding author.

## Acknowledgements

NSF#2033307-PI CA, intramural support from Plexision.

